# COVID-19 pandemic reshaped seasonal patterns and age distributions of respiratory viruses

**DOI:** 10.1101/2024.12.25.24319311

**Authors:** Miao He, Lanping Gao, Yanwu Hu, Simeng Liang, Jiajia Tang, Yongliang Zhao, Youwei Zhang, Na Deng, Chanjuan Zhou, Haoyue Wang, Xiyu Lan, Zixuan Wang, Yanran Li, Ke Xu

**Author notes:** These authors contributed equally to this work (Miao He, Lanping Gao, Yanwu Hu, Simeng Liang, Jiajia Tang). Corresponding authors. Address correspondence and reprint requests to Dr. Ke Xu), and Dr. Miao He).

## Abstract

**Objectives:** This study aims to investigate the changes in the prevalence and demographic characteristics of common respiratory viruses during and after the COVID-19 pandemic.

**Methods:** We retrospectively enrolled children with acute respiratory infections (ARIs) at Shiyan Renmin Hospital, Hubei University of Medicine, from January 2020 to December 2023. Specimens serum, nasopharyngeal aspirate, and alveolar lavage fluid were collected for direct immunofluorescence assay (DFA): respiratory syncytial virus (RSV), adenovirus (ADV), influenza A virus (IAV), influenza B virus (IBV), and parainfluenza virus (PIV). Demographic data and laboratory test results were analyzed accordingly.

**Results:** A total of 10,193 patients were enrolled. The positive infection rates for the years 2020, 2021, 2022, and 2023 were 3.97%, 3.15%, 36.20%, and 38.82%, respectively. The seasonal patterns for ADV transitioned from peaking in the summer and autumn of 2022 to summer and winter in 2023, while RSV peaked in the spring and summer of 2022 but moved to spring and autumn in 2023. PIV shifted from autumn 2022 to both spring and autumn in 2023. Intriguingly, IAV stably remained a two-season pattern of summer and winter in 2022 and 2023, while IBV showed up at 2022 winter but largely diminished later. The age distribution of children infected with ADV, RSV and PIV showed an upward trend, while no significant changes were observed for IAV and IBV.

**Conclusions:** The COVID-19 pandemic has disrupted the seasonal circulation of respiratory viruses. The seasonal pattern of influenza virus has been restored in 2022. In contrast, ADV, PIV, and RSV showed significant seasonal changes after the pandemic. The increasing age distributions of cases indicates an expanded age range of infection. Continuous monitoring of pathogen distribution and adjustment of preventive strategies are crucial for the effective management of pediatric ARIs.

## Introduction

Since the emergence of coronavirus disease 2019 (COVID-19) at the end of 2019, the virus has rapidly spread across numerous countries and regions globally.^1^ The COVID-19 pandemic has profoundly reshaped the global health landscape, impacting various aspects of public health, including the epidemiology of acute respiratory infections (ARIs) in children.^2,3^ Traditional surveillance systems and established epidemiological patterns have been disrupted due to the implementation of non-pharmaceutical interventions (NPIs), such as school closures, social distancing, and mask mandates.^4–6^ While these measures have proven effective in mitigating the transmission of SARS-CoV-2, they have also impacted the incidence and prevalence of other respiratory pathogens.^7,8^

Previous studies have demonstrated a decrease in the incidence of influenza and other respiratory syncytial virus (RSV) infections during the early phases of the pandemic, which has been attributed to the widespread implementation of NPIs.^9–11^ However, as the NPIs restrictions were relaxed, a resurgence of such infections was observed, leading to the emergence of the concepts of “immunity debt” or “immunity gap”.^12^ This suggests that the limited exposure to pathogens during the pandemic might have resulted in a decline in overall population immunity, making the susceptible population more sensitive to respiratory infections. Local hospitalized children are among these susceptible populations, highlighting the necessity of surveilling the epidemiological characteristics of acute respiratory pathogens for developing effective public health policies and preventive control measures. In addition, testing for respiratory viral pathogens is an important guide for the early identification of the type of viral infection, the selection of antiviral drugs, and the prevention of drug abuse.

This study aims to provide a comprehensive epidemiological analysis of ARIs in children during the COVID-19 pandemic, focusing on temporal trends, age distribution, and geographical variations from a single hospital location. By leveraging data from pediatric hospitals and public health surveillance systems, we seek to understand the interplay between NPIs, viral circulation, and the health outcomes of children. We analyze the epidemiological characteristics of respiratory viruses such as Adenovirus (ADV), RSV, Influenza A Virus (IAV), Influenza B Virus (IBV), and Parainfluenza Virus (PIV) in ARIs among hospitalized children during and post the COVID-19 pandemic in Shiyan, Hubei Province.

With over 3.5 million residents and significant population mobility as a regional transit hub, the city Shiyan offers a unique setting to study IBV, PIV, and ARIs in children, reflecting both local and regional epidemiological trends. The findings of this research will contribute to the development of evidence-based strategies for managing ARIs in the post-pandemic era, ensuring the protection of vulnerable pediatric populations.

## Methods

### Study Design and Participants

This study included a total of 10,193 children under the age of 16 years who were hospitalized with signs of ARI at the Department of Pediatrics at Shiyan Renmin Hospital, Hubei University of Medicine, from January 2020 to December 2023. The inclusion criteria for the diagnosis of acute respiratory infection were as follows: If the patient has (1) at least one manifestation of acute infection, including fever (≥ 38°C), abnormal white blood cell (WBC) counts (leukocytosis with WBC count > 10,000/ml or leukopenia with WBC count < 4,000/ml), and chills, in conjunction with at least one respiratory symptom such as pharyngeal discomfort, dry throat or sore throat, nasal congestion, rhinorrhea, significant congestion and edema of the nose, pharynx, and larynx, cough, expectoration, or shortness of breath; (2) the presence of abnormal breath sounds upon auscultation (moist rales, dry rales, wheezing, and dullness) and chest pain. The exclusion criteria included congenital heart disease, immunocompromised status, allergic rhinitis, tuberculosis, bronchial asthma, and cases with incomplete clinical data.

Numerical clinical data, including demographic, epidemiological, diagnostic, and laboratory information, were recorded via the hospital’s electronic medical record system. Meanwhile, research subjects with incomplete medical records were excluded to ensure the representativeness and homogeneity of the research samples.

The study was approved by the Ethics Committee of Shiyan Renmin Hospital, Hubei University of Medicine (File NO. SYRMYY-2024-137). Written consent was waived because antigen detection of respiratory pathogens is standard and routine for all children admitted for ARIs in the hospital while the study was implemented.

### Data and Specimen Collection

Specimens, including serum, nasopharyngeal aspirate, and alveolar lavage fluid, were collected from all enrolled children within 72 hours of their admission by trained personnel, adhering to established standard operating procedures. Demographic and clinical data for the enrolled children were obtained from their electronic medical records.

### Specimen Detection

Five respiratory viruses, namely ADV, RSV, IAV, IBV, and PIV, were detected using an immune rapid antigen detection kit (PNEUMOSLIDE IgM, Bilthai Co., Ltd., VIRCELL, S.L., Spain) in 2020-2021, during 2022-2023, a five respiratory pathogen nucleic acid test kit (Guangdong Hexin Health Technology Co., Ltd.) was used for detection.

### Statistical Analysis

Summary statistics for categorical variables were presented as frequencies (percentages), while means, medians, or interquartile ranges (IQR) were used for continuous variables. Comparisons of categorical variables were performed using chi-squared tests or Fisher’s exact tests, and t-test was used for the comparison between the groups. All statistical tests were two-tailed, and p-values less than 0.05 were considered statistically significant. Statistical analyses were conducted using SPSS version 26.0 (IBM Corp, Armonk, NY, USA).

## Results

### Demographic characteristics of study population

To investigate the changes in the demographic characteristics of respiratory viruses during and after the COVID-19 pandemic, we analyzed data from a total of 10,193 hospitalized children with ARI recorded between January 2020 and December 2023 tested for five respiratory viruses. The characteristics of the children enrolled in this study are summarized in Table 1. Among these children, 5,702 (55.94%) were male and 4,491 (44.91%) were female and there were no significant differences detected in the positive rate of respiratory virus infections by gender (p > 0.05) (Supplementary Table 1). The age of participants ranged from 1 day to 16 years, with a median age of 39.87 months (IQR: 14.1-68.6 months). The frequency within age cohorts of < 31 days, >31 days - <1 year, 1-3 years, 4-6 years, 7-12 years, and >12 years were 1.74%, 20.20%, 23.87%, 31.43%, 21.58%, and 1.17%, respectively. Notably the positive rate was significantly higher in the 7-12 years age group compared to the other age groups (p < 0.0001). A majority of patients (77.25%) with ARI were under 6 years old. According to the temperature-associated seasonal fluctuations, the patients were divided into four cohorts: (1) Spring (March–May), 2,295 (22.52%); (2) Summer (June–August), 2,164 (21.23%); (3) Autumn (September–November), 3,033 (29.76%); (4) Winter (December–February), 2,701 (26.50%). Saliently, the positive rate in summer group was significantly higher compared to the other seasons (p < 0.0001) (Supplementary Table 1)

**Table 1:** Demographic characteristics of participants (n=10,193) in the study.

| Variable | Category | Total (n=10,193), n (%) |
| --- | --- | --- |
| Gender | Male | 5,702(55.94) |
|  | Female | 4,491(44.06) |
| Age group | ≤ 31 days | 178(1.74) |
|  | > 31days-< 1year | 2,059(20.20) |
|  | 1–3 years | 2,433(23.87) |
|  | 4–6 years | 3,204(31.43) |
|  | 7–12years | 2,200(21.58) |
|  | >12 years | 119(1.17) |
| Season | Spring | 2,295(22.52) |
|  | Summer | 2,164(21.23) |
|  | Autumn | 3,033(29.76) |
|  | Winter | 2,701(26.50) |

### Annual changes in the prevalence of respiratory viruses

We further detected the positive rates of five respiratory viruses in hospitalized children (2020–2023). The annual test numbers and positive rates from 2020 to 2023 were as follows: 1,434 cases (3.97%, 57/1,434) in 2020; 1,111 cases (3.15%, 35/1,111) in 2021, with the lowest number of tests and positivity; 1,870 cases (36.20%, 677/1,870) in 2022; and 5,778 cases (38.82%, 2,243/5,778) in 2023 (Figure. 1A). In 2020 and 2021, the monthly detection numbers, positive cases, and positive rates decreased sharply, particularly from January 23 to April 8, 2020, coinciding with the blockade period, and remained at relatively low levels thereafter. An increase occurred only at the start of the new semester in September 2020, peaking again in winter 2020. In 2022 and 2023, an upward trend was observed, with peaks in spring and summer months (Figure. 1B).

**Figure 1.**
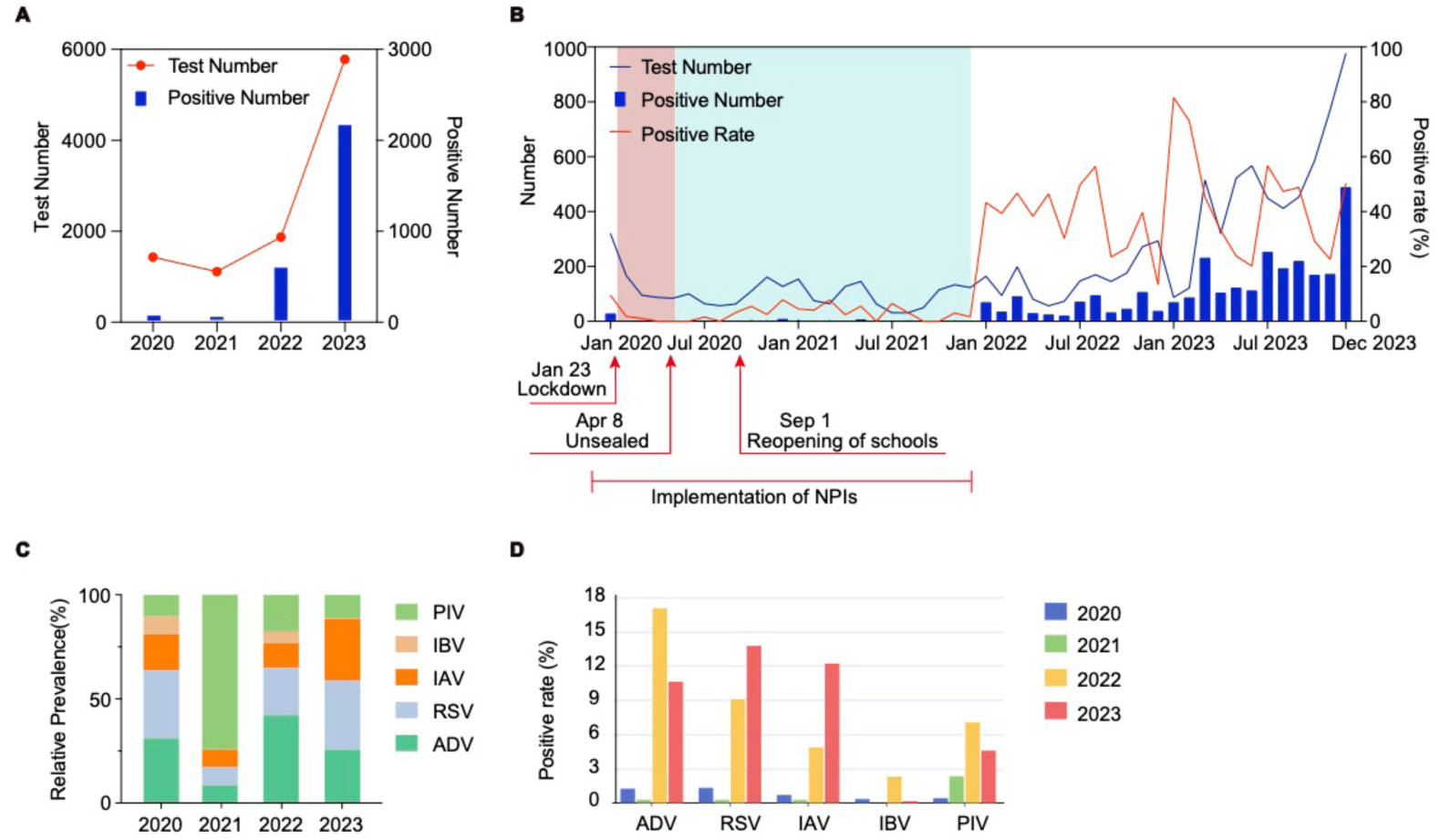
Distribution and positive rate of 10,193 hospitalized children with ARIs from January 2020 to December 2023. (A) Total number of tested and number of positive tested per year. (B) Monthly distribution and positive rate of respiratory viruses. The lockdown period from January 23 to April 8, 2020 is shown in pink, and the post-lockdown period is shown in blue. (C) Relative prevalence of each respiratory virus by year. (D) Positive rates by virus type and year. Abbreviations: ADV, adenovirus; IAV, influenza A virus; IBV, influenza B virus; PIV, parainfluenza virus; RSV, respiratory syncytial virus; NPIs, non-pharmaceutical interventions.

The five viruses showed different epidemiological patterns over a 4-year period, with their relative positive rates from 2020 to 2023 detailed in Figure. 1C: ADV and RSV were predominant in 2020; PIV was predominant in 2021, with similar positive rates of ADV, RSV, and IAV, while IBV was absent throughout the year. In 2022, as NPIs were gradually relaxed, ADV became the predominant circulating virus, the relative positivity of PIV declined compare to 2021 and IBV re-emerged. In 2023, with the epidemic fully relaxed, RSV replaced ADV as the predominant virus, and the relative positivity of IAV was elevated while IBV was significantly reduced.

We compared the change in positive rates of each virus from 2020 to 2023. In 2022, the positive rate of ADV (17.06%, 319/1,870) was significantly higher than in 2020 (18/1,434, 1.25%), 2021 (0.27%, 3/1,111), and 2023 (10.63%, 614/5,778). In 2023, the positive rate of RSV (13.78%, 796/5,778) was significantly higher than in 2020 (1.32%, 19/1,434), 2021 (0.27%, 3/1,111), and 2022 (9.09%, 170/1,870). IAV had the highest positive rate in 2023 (12.22%, 706/5,778), followed by 2022 (4.87%, 91/1,870), 2020 (0.7%, 10/1,434), and 2021 (0.27%, 3/1,111). The IBV positive rate was highest in 2022 (2.30%, 43/1,870), followed by 2020 (0.35%, 5/1,434) and 2023 (0.16%, 9/5,778). For PIV, the positive rate in 2022 was significantly higher than in 2020 (0.42%, 6/1,434), 2021 (2.34%, 26/1,111), and 2023 (4.59%, 265/5,778) (Figure. 1D and Supplementary Table 2). The circulations of five respiratory viruses rebounded in 2022, though the dominant virus remained in flux.

### Seasonal changes in the prevalence of respiratory viruses

We further analyzed the monthly and seasonal dynamic patterns of five respiratory viruses from 2020 to 2023. In 2020-2021, the positivity rate for all five viruses was extremely low and did not show seasonality. In contrast, these five viruses rebounded in 2022 and 2023, although with abnormal seasonality. ADV was detected throughout 2022 and 2023 with the annual peak for ADV occurring in May 2022 (37.5%, 21/56), and in July 2023 (30.07%, 135/449) (Figure. 2A). It is noteworthy that PIV was detected throughout 2022, with peak activities in March (15.58%, 31/199), January (14.63%, 24/164) and July (14.97%, 22/147). In 2023, PIV showed peak activity in April (20.94%, 67/320) and October (13.7%, 80/584) (Figure. 2B). For RSV, the highest positive rate in 2022 appeared in November (22.43%, 61/272), while the highest positive rate in 2023 occurred in March (34.95%, 180/515), with a peak in September 2023 (34.66%, 157/453) (Figure. 2C). For IAV, it was detected only starting from May 2022, with the highest positive rate for the year occurring in August (36.47%, 62/170). In 2023, the highest annual positive rate occurred in February (67.21%, 82/122), and another peak emerged in August (43.45%, 179/412) (Figure. 2D). In 2022, IBV was predominantly prevalent from January to March, with the highest positive rate in March (9.05%, 18/199) and sporadic occurrences in the other months. In 2023, IBV was nearly diminished throughout the whole year (Figure. 2E).

**Figure 2.**
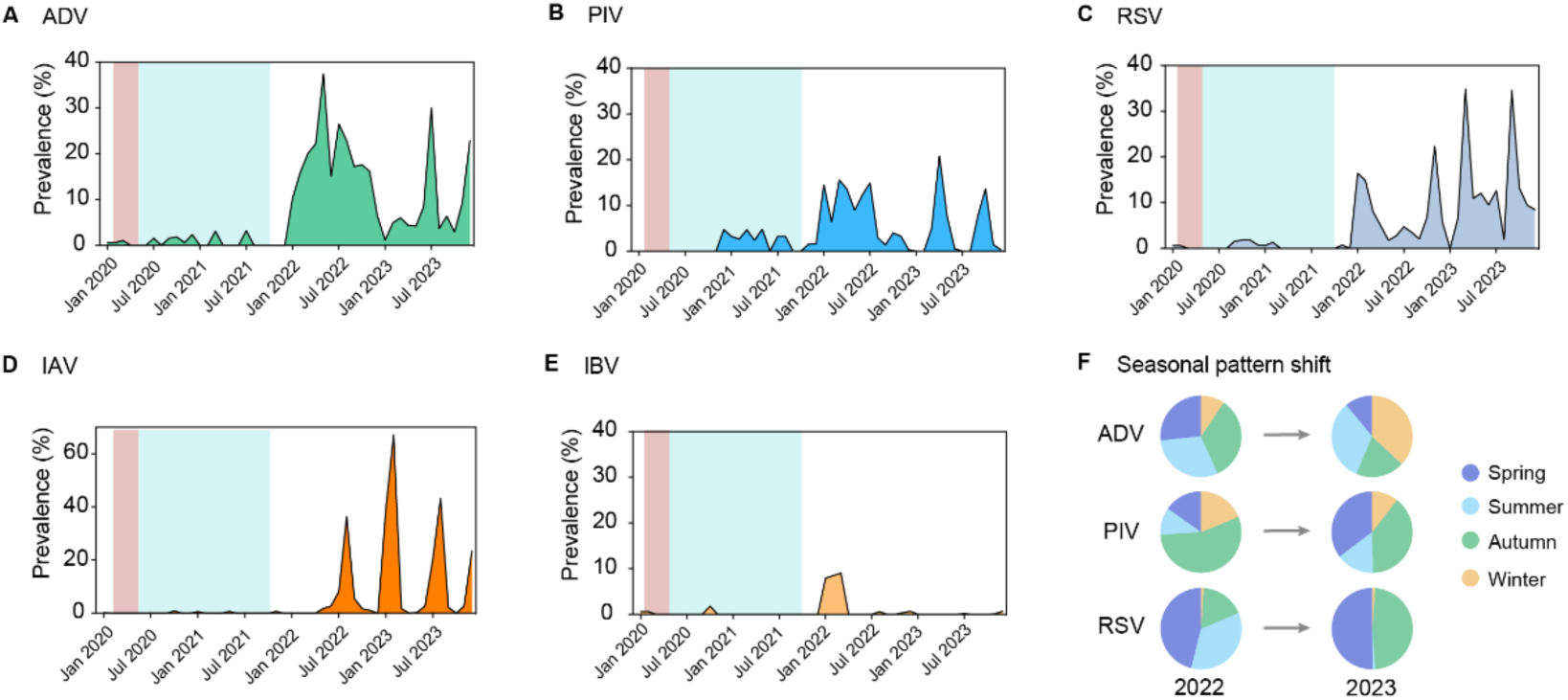
Monthly distribution and positive rate of specific respiratory viruses among 10,193 hospitalized children from January 2020 to December 2023. (A-E) Monthly distribution and positive rate of ADV (A), PIV (B), RSV (C), IAV (D), and IBV (E). (F) Seasonal activity of respiratory viruses in 2022 and 2023.

Meanwhile, we found that ADV, PIV, and RSV exhibited distinct seasonal distributions between 2022 and 2023. ADV was prevalent in spring, summer, and autumn in 2022, with the highest positive rate in autumn. In 2023, it was concentrated in summer and winter, peaking in winter. For PIV, it was prevalent in spring and summer in 2022, with the highest positive rate in spring. In 2023, the highest positive rate was also in spring, with another peak in autumn. RSV was mainly prevalent in autumn in 2022, and the positive rate in 2023 was also in autumn, but also peaked in spring (Figure. 2F, supplementary Table 3). IAV was prevalent in summer and winter in both 2022 and 2023, while the positive rate of IBV was too low to be representative.

### The trend of increasing age among infected children

The five viruses exhibited different distribution patterns across six age groups. Among the distribution of positive cases for the five viruses, IAV had the highest median age (5 years and 3 months), followed by ADV (4 years and 7 months), IBV (4 years), RSV (2 years and 9 months), and PIV, which had the youngest median age (2 years and 8 months) (Figure. 3A). The positive rate of IAV was highest in the 7-12 years old group (10.21%, 226/2,200). ADV had the highest positive rate in the over-12-year-old group (11.76%, 14/119). The positive rate of IBV was highest in the neonatal group (1.12%, 2/178) and was not detected in the 7-12 years group. The RSV positive rate increased with age and peaked in the 7-12-year-old group (12.59%, 227/2,200). PIV had the highest positivity rate in the 4-6-year-old cohort (4.81%, 154/3,204) (Supplementary Table 4).

**Figure. 3.**
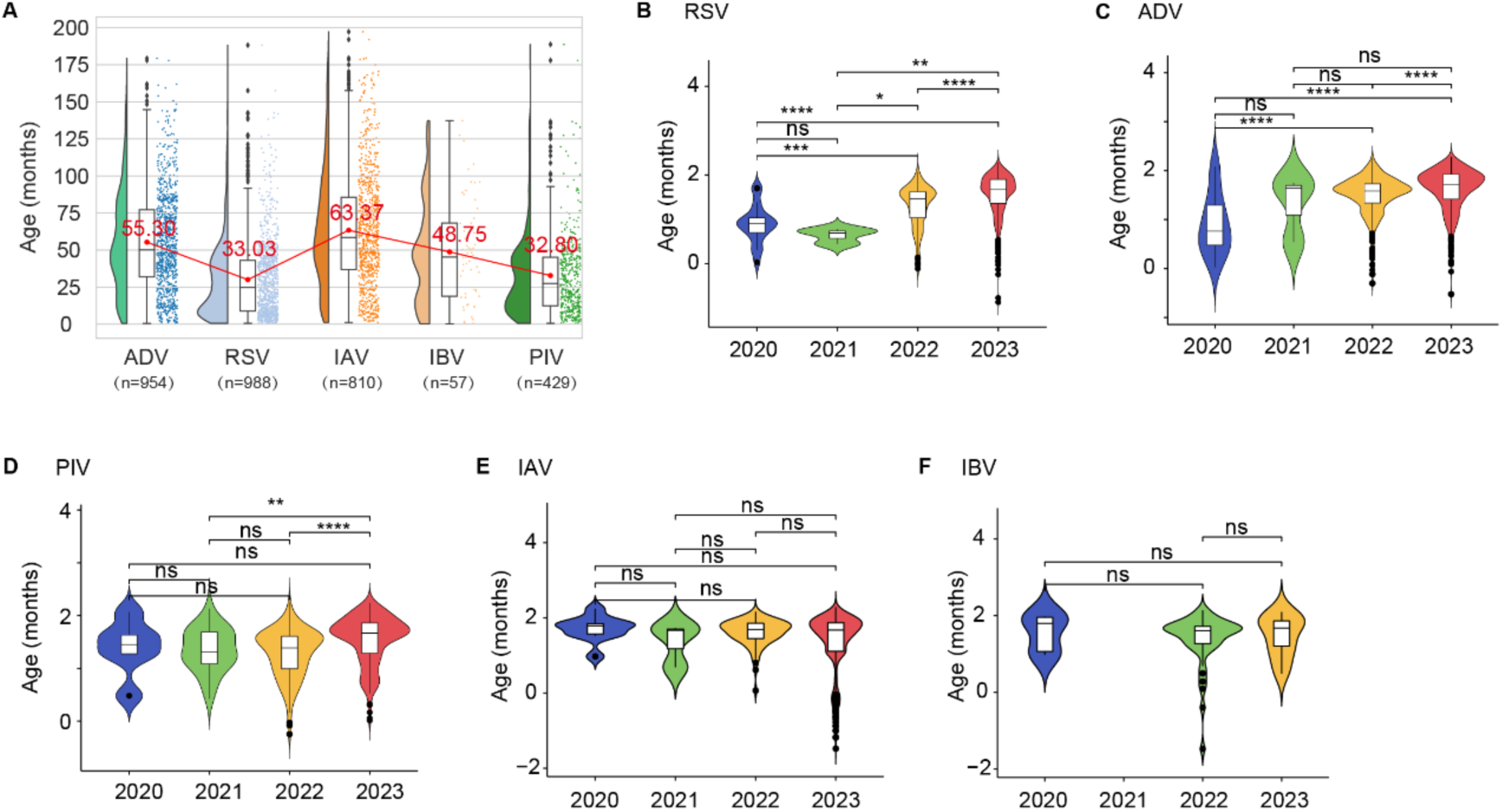
The distribution of virus-positive cases across age groups and years. (A) Age spectrum of respiratory viruses. (B-F) The age distribution of the population infected with RSV (B), ADV (C), PIV (D), IAV (E), and IBV (F) each year. Superscripts like ‘*’ are p-values of the t-test, ‘ns’, p-value > 0.05, ‘*’, p value < 0.05, ‘**’, p-value < 0.01, ‘***’, p-value <0.001, ‘****’, p-value < 0.0001.

Additionally, we found that RSV, ADV, and PIV exhibited significant age fluctuations over time. Among them, RSV showed an increasing trend in the age of susceptible individuals from 2020 to 2023. Although the median decreased from 2020 to 2021, the difference was not statistically significant (Figure. 3B). ADV had the highest median age in 2021 with no significant difference with 2023. In 2023, there was a significant increase in the age of the susceptible population for ADV compared to 2020 and 2022 (Figure. 3C). For PIV, the age distribution of the susceptible population was higher in 2023 compared to 2022 and 2021 (Figure. 3D). In contrast, for IAV and IBV, whose age distributions exhibited no significant fluctuations in different years (Figure. 3E and 3F).

## Discussion

This study presents insights into the epidemiological patterns and demographic changes of respiratory viruses among hospitalized children with ARIs at Shiyan Renmin Hospital, Hubei University of Medicine, from January 2020 to December 2023. Notable shifts were observed in the prevalence, seasonality, and age distribution of RSV, ADV, and PIV during and after the COVID-19 pandemic. In contrast, the seasonal patterns and age distributions of influenza remained stable in 2022 and 2023, highlighting the influenza virus dynamics returned to pre-pandemic levels. These findings underscore the lasting impacts of the COVID-19 pandemic on these respiratory viral dynamics.

The implementation of stringent NPIs during COVID-19 pandemic significantly reduced the number of respiratory virus infections, as reflected by low positive test rates in this study. Similarly, other studies have reported a decline in pediatric hospitalizations for infectious diseases during this period.^13–19^ Among these viruses, the influenza virus experienced a notable suppression due to NPIs from 2020 to 2021.^20^ However, the WHO’s pandemic influenza intervention guidance does not recommend stringent NPIs for management of influenza pandemics.^21^ Thus, we suggest a need to re-evaluate the role of stringent NPIs in mitigating or even eliminating severe pandemic influenza. As COVID-19 pandemic was under control and NPIs were gradually relaxed, the respiratory viral positivity rate rose sharply in 2022 and 2023, with a pronounced rebound during the easing of restrictions in 2023 under Category B control.^22^ The prevalence and seasonality of RSV, ADV, and PIV from 2022 to 2023 continued to exhibit shifts. These findings inform better assessment of NPIs on delaying, containing or averting transmission for respiratory viral threats and future pandemics.

The resurgence of non-SARS-CoV-2 viruses likely resulted from waning population immunity due to prolonged reduced exposure to pathogens, as posited by the “immune liability” hypothesis.^23,24^ This theory suggests that strict NPIs disrupt the regular stimulation and maintenance of herd immunity by significantly reducing pathogen circulation. As a result, populations become increasingly susceptible to infections once restrictions are lifted, creating the conditions for accelerated transmission and larger outbreaks. The rebound effect observed in this study further highlights the interplay between herd immunity and viral transmission dynamics. Herd immunity is achieved when a significant portion of the population develops immunity, either through infection or vaccination, thereby reducing the likelihood of transmission and protecting those who are not immune.^25,26^ However, during periods of strict NPIs, vaccination against respiratory viruses became an essential tool to maintain protection among susceptible hosts, as natural exposure was minimized. This highlights the need for encouraging vaccination of susceptible individuals to sustain long-term immunity within populations.

Further, we observed an increase in the median age of children infected with ADV, RSV, and PIV from 2022 to 2023, consistent with findings from other studies. For instance, an unusual RSV outbreak in 2022-2023 showed hospitalization rates surge among children older than the typical peak-risk age group.^27^ Similarly, a report from Australia highlighted an upward shift in the age distribution of PIV cases during the post-pandemic rebound, with older children experiencing increased clinical severity.^15^ These shifts likely result from delayed primary infections in younger children due to reduced exposure during the COVID-19 pandemic. These demographic changes may influence the clinical presentation and healthcare burden of these infections. This evolving age distribution underscores the need for tailored preventive strategies, such as adjusting vaccination schedules.

While this study offers valuable insights, certain limitations should be acknowledged. First, these findings are based on data collected by direct immunofluorescence assays (DFA), which might limit the detection of less prevalent respiratory pathogens. Future studies incorporating multicenter data and advanced diagnostic techniques, such as multiplex PCR, could provide a more comprehensive understanding of respiratory virus dynamics.^28,29^ Additionally, the present study focused exclusively on common acute respiratory viruses but excluded bacterial pathogens, resulting in an incomplete representation of the spectrum of infectious agents associated with ARIs.^30^ Future research should expand to include bacterial testing and leverage multicenter data to improve generalizability.

In summary, the COVID-19 pandemic has profoundly influenced the epidemiology of respiratory viruses in children, with significant alterations in their prevalence, seasonal patterns, and age distribution. This study underscores the necessity for continuous surveillance of respiratory pathogens to detect and respond to changes in their epidemiology. Targeted public health interventions including vaccination strategies, diagnostic protocols, and treatment guidelines are critical to mitigate the burden of pediatric ARIs in this dynamic post-pandemic environment.

## Conclusions

In this study, more than 10,000 hospitalized children diagnosed with respiratory tract infections during and after the COVID-19 epidemic were enrolled to explore the prevalence of respiratory viruses. The results showed that non-pharmacological interventions implemented during the COVID-19 pandemic had a significant impact on the prevalence and seasonal patterns of common respiratory viruses. We should recognize that seasonal respiratory viruses may not present typical epidemiological patterns and be cautious of a possible that unusual rebound and unexpected resurgence when health restrictions are fully lifted. Therefore, multiple factors should be taken into account when predicting the epidemiological trends of pathogens, and continuous monitoring of the epidemiological and evolutionary dynamics of a wide range of respiratory pathogens is essential.

## Author Contributions

K.X. conceived the project and designed the experiments. M.H., LP.G., YW.H, and JJ.T. collected and analyzed clinical data. SM. L., YL.Z., YW.Z., N.D., CJ.Z., HY.W., XY.L., ZX.W., and YR.L. provided clinical data and information and contributed to data interpretation. K.X., M.H., LP.G., and SM. L. wrote the manuscript with input from all the other authors. All authors approved the final manuscript.

## Data Availability

All data produced in the present study are available upon reasonable request to the authors

