## Supplemental Table 1-4 for "COVID-19 pandemic reshaped seasonal patterns and age distributions of respiratory viruses"

| Supplementary Table 1: Distribution of positive cases in the study | | | | |
| --- | --- | --- | --- | --- |
| Variable | Category | Total (n=10,193), n (%) | χ2 | p |
| Gender | Male | 1,658 (29.08) | 1.386^a^ | 0.239 |
|  | Female | 1,354 (30.15) |  |  |
| Age of child (years) | ≤31 days | 19 (10.67) | 47.036^a^ | <0.001*** |
|  | > 31days– < 1year | 500 (24.28) |  |  |
|  | 1–3 years | 693 (28.48) |  |  |
|  | 4–6 years | 1,025 (31.20) |  |  |
|  | 7–12years | 739 (33.60) |  |  |
|  | >12 years | 36 (30.26) |  |  |
|  | Spring  Summer  Autumn  Winter | 630 (27.45)  759 (35.07)  731 (25.42)  852 (31.54) | 66.580^a^ <0.001*** | |
| Season |  |  |  |  |

* p < 0.05 , ** p < 0.01, *** p < 0.001

| Supplementary Table 2: Distribution of respiratory viruses by year | | | | |
| --- | --- | --- | --- | --- |
| Category | Year | n, n(%) | χ2 | p |
| ADV | 2020^b^ | 18, 1.25 | 360.825^a^ | 0.000 |
|  | 2021^c^ | 3, 0.27 |  |  |
|  | 2022^d^ | 319, 17.06 |  |  |
|  | 2023^e^ | 614, 10.63 |  |  |
| RSV | 2020^b^ | 19, 1.32 | 338.250^a^ | 0.000 |
|  | 2021^c^ | 3, 0.27 |  |  |
|  | 2022^d^ | 170, 9.09 |  |  |
|  | 2023^e^ | 796, 13.78 |  |  |
| IAV | 2020^b^ | 10, 0.7 | 360.935^a^ | 0.000 |
|  | 2021^b^ | 3, 0.27 |  |  |
|  | 2022^c^ | 91, 4.87 |  |  |
|  | 2023^d^ | 706, 12.22 |  |  |
| IBV | 2020^b^ | 5, 0.35 | 126.146^a^ | 0.000 |
|  | 2021^b^ | 0, 0 |  |  |
|  | 2022^c^ | 43, 2.30 |  |  |
|  | 2023^b^ | 9, 0.16 |  |  |
| PIV | 2020^b^ | 6, 0.42 | 100.442^a^ | 0.000 |
|  | 2021^c^ | 26, 2.34 |  |  |
|  | 2022^d^ | 132, 7.06 |  |  |
|  | 2023^e^ | 265, 4.59 |  |  |

Superscript letters like “b, c, d, e” on years represent pairwise comparisons of virus positive rates. Different letters mean statistically significant differences (p < 0.05), while the same letters indicate the opposite.

| Supplementary Table 3: Seasonal distribution of respiratory viruses by year | | | | | | |
| --- | --- | --- | --- | --- | --- | --- |
| Category | Number of case | ADV(%) | RSV(%) | IAV(%) | IBV(%) | PIV(%) |
| 2020 | Spring(n=266) | 1(0.08) | 0(0) | 0(0) | 0(0) | 0(0) |
|  | Summer(n=221) | 1(0.08) | 0(0) | 0(0) | 0(0) | 0(0) |
|  | Autumn(n=334) | 4(0.34) | 6(0.51) | 1(0.08) | 2(0.17) | 0(0) |
|  | Winter(n=356) | 3(0.25) | 3(0.25) | 1(0.08) | 0(0) | 13(1.10) |
|  | χ2 | 1.667^a^ | 8.467^a^ | 1.417^a^ | 5.056^a^ | 30.315^a^ |
|  | p | 0.644 | 0.037 | 0.701 | 0.168 | 0.000 |
| 2021 | Spring(n=336) | 2(0.18) | 0(0) | 1(0.09) | 0(0) | 13(1.14) |
|  | Summer(n=125) | 1(0.09) | 0(0) | 0(0) | 0(0) | 2(0.18) |
|  | Autumn(n=298) | 0(0) | 1(0.09) | 1(0.09) | 0(0) | 2(0.18) |
|  | Winter(n=381) | 32(2.81) | 41(3.60) | 0(0) | 21(1.84) | 32(2.81) |
|  | χ2 | 54.874^a^ | 80.825^a^ | 1.612^a^ | 42.620^a^ | 27.469^a^ |
|  | p | 0.000 | 0.000 | 0.657 | 0.000 | 0.000 |
| 2022 | Spring(n=336) | 79(4.34) | 21(1.15) | 1(0.05) | 18(0.99) | 47(2.59) |
|  | Summer(n=390) | 89(4.89) | 15(0.82) | 76(4.17) | 1(0.05) | 36(1.98) |
|  | Autumn(n=593) | 100(5.50) | 76(4.17) | 14(0.77) | 1(0.05) | 18(0.99) |
|  | Winter(n=502) | 28(2.54) | 26(1.43) | 150(8.24) | 2(0.11) | 1(0.05) |
|  | χ2 | 67.552^a^ | 35.956^a^ | 244.443^a^ | 59.547^a^ | 89.502^a^ |
|  | p | 0.000 | 0.000 | 0.000 | 0.000 | 0.000 |
| 2023 | Spring(n=1357) | 67(1.20) | 278(5.00) | 11(0.20) | 0(0) | 133(2.39) |
|  | Summer(n=1428) | 198(3.56) | 119(2.14) | 285(5.12) | 1(0.02) | 3(0.05) |
|  | Autumn(n=1808) | 117(2.10) | 307(5.51) | 31(0.56) | 1(0.02) | 126(2.26) |
|  | Winter(n=976) | 223(4.00) | 83(1.50) | 229(4.11) | 7(0.13) | 3(0.05) |
|  | χ2 | 243.296^a^ | 122.284^a^ | 620.017^a^ | 22.878^a^ | 203.481^a^ |
|  | p | 0.000 | 0.000 | 0.000 | 0.000 | 0.000 |

| Supplementary Table 4: Distribution of respiratory viruses in gender and age | | | | | | | | | | | | | | | | |
| --- | --- | --- | --- | --- | --- | --- | --- | --- | --- | --- | --- | --- | --- | --- | --- | --- |
| Variables | Category | ADV | | | RSV | | IAV | | | IBV | | | PIV | | | |
|  |  | The positive rate (n/n) | χ2 | p | The positive rate (n/n) | χ2 | p | The positive rate (n/n) | χ2 | p | The positive rate (n/n) | χ2 | p | The positive rate (n/n) | χ2 | p |
| Gender | Male | 9.26(528/5,702) | 1.152^a^ | 0.283 | 9.24(527/5,702) | 3.001^a^ | 0.083 | 7.96(454/5,702) | 0.004^a^ | 0.948 | 0.49(28/5,702) | 1.081^a^ | 0.298 | 4.40(251/5,702) | 1.198^a^ | 0.274 |
|  | Female | 9.71(436/4,491) |  |  | 10.26(461/4,491) |  |  | 7.93(356/4,491) |  |  | 0.65(29/4,491) |  |  | 3.96(178/4,491) |  |  |
| Age of  child (years) | ≤31 days | 2.81(5/178) |  | | 3.93(7/178) |  |  | 2.25(4/178) |  | | 1.12(2/178) |  | | 1.69(3/178) |  | |
|  | >31days–<1year | 6.31(130/2,059) | 47.036^a^ | <0.001*** | 8.31(171/2,059) | 15.482^a^ | 0.008*** | 6.41(132/2,059) | 32.499^a^ | <0.001*** | 0.91(9/2,059) | 3.621^a^ | 0.605 | 4.37(90/2,059) | 9.686^a^ | 0.085 |
|  | 1–3 years | 9.08(221/2,433) |  | | 9.49(231/2,433) |  |  | 7.64(186/2,433) |  | | 0.44(14/2,433) |  | | 4.23(103/2,433) |  | |
|  | 4–6 years | 10.46(335/3,204) |  | | 10.61(340/3,204) |  |  | 7.99(256/3,204) |  | | 0.69(22/3,204) |  | | 4.81(154/3,204) |  | |
|  | 7–12years | 11.32(249/2,200) |  | | 12.59(227/2,200) |  |  | 10.27(226/2,200) |  | | 0.45(10/2,200) |  | | 3.36(74/2,200) |  | |
|  | >12 years | 11.76(14/119) |  | | 10.08(12/119) |  |  | 5.04(6/119) |  | | 0(0) |  | | 4.20(5/119) |  | |

* p<0.05, ** p<0.01 , *** p<0.001
